# Bidirectional relationship between sleep problems and long COVID: a longitudinal analysis of data from the COVIDENCE UK study

**DOI:** 10.1101/2024.02.08.24302486

**Authors:** Giulia Vivaldi, Mohammad Talaei, John Blaikley, Callum Jackson, Paul E Pfeffer, Seif O Shaheen, Adrian R Martineau

## Abstract

**Background:** Studies into the bidirectional relationship between sleep and long COVID have been limited by retrospective pre-infection sleep data and infrequent post-infection follow-up. We therefore used prospectively collected monthly data to evaluate how pre-infection sleep characteristics affect risk of long COVID, and to track changes in sleep duration during the year after SARS-CoV-2 infection.

**Methods:** COVIDENCE UK is a prospective, population-based UK study of COVID-19 in adults. We included non-hospitalised participants with evidence of SARS-CoV-2 infection, and estimated odds ratios (ORs) for the association between pre-infection sleep characteristics and long COVID using logistic regression, adjusting for potential confounders. We assessed changes in sleep duration after infection using multilevel mixed models. We defined long COVID as unresolved symptoms at least 12 weeks after infection. We defined sleep quality according to age-dependent combinations of sleep duration and efficiency. COVIDENCE UK is registered with ClinicalTrials.gov, NCT04330599.

**Findings:** We included 3994 participants in our long COVID risk analysis, of whom 327 (8.2%) reported long COVID. We found an inverse relationship between pre-infection sleep quality and risk of long COVID (medium *vs* good quality: OR 1.37 [95% CI 1.04–1.81]; medium–low *vs* good: 1.55 [1.12–2.16]; low *vs* good: 1.94 [1.11–3.38]). Greater variability in pre-infection sleep efficiency was also associated with long COVID (OR per percentage-point increase 1.06 [1.01–1.11]). We assessed post-infection sleep duration in 6860 participants, observing a 0.11 h (95% CI 0.08–0.13) increase in the first month after infection compared with pre-infection, with larger increases for more severe infections. After 1 month, sleep duration largely returned to pre-infection levels, although fluctuations in duration lasted up to 6 months after infection among people reporting long COVID.

**Interpretation:** Our findings highlight the bidirectional relationship between sleep and long COVID. While poor-quality sleep before SARS-CoV-2 infection associates with increased risk of long COVID thereafter, changes in sleep duration after infection in these non-hospitalised cases were modest and generally quick to resolve.

**Funding:** Barts Charity.

## Introduction

Long COVID, which describes the post-acute sequelae of SARS-CoV-2 infection, is estimated to affect at least 10% of people who are infected.^1^ One of the most common symptoms of long COVID is poor sleep quality, which is reported by an estimated 45% of COVID-19 survivors,^2^ with higher prevalence among those with severe disease.^3^ However, different timings of assessment across studies and infrequent follow-up has made it difficult to track the trajectory of sleep after infection,^4^ with the majority of data only covering the first 6 months after infection.^2^ Additionally, few studies have factored in pre-infection sleep,^2,5^ and those that do often rely on retrospectively collected data, which is subject to recall bias and can be unreliable.^6^ The situation is further complicated by the economic and public health restrictions present during the COVID-19 pandemic, during which stress, restrictions to movement, loss of morning commutes, and changes to employment are likely to have increased or decreased sleep duration and quality at different timepoints, independently of infection.^6,7^

Importantly, the relationship between sleep and long COVID may be bidirectional. Pre-existing sleep problems have been shown to increase the risk of upper respiratory infections, including influenza and SARS-CoV-2, as well as hospitalisation with COVID-19.^8^ Sleep quality has also been implicated in the development of long COVID among those infected, with studies finding a wide range of sleep-related measures such as sleep quality, sleep duration, and pre-existing sleep disorders predicting the development of long COVID.^9–12^ This research, however, is still in its infancy, with studies presenting conflicting findings on the associations with different sleep measures;^9,10^ heterogeneity between study definitions of long COVID, ranging between symptoms lasting 4 weeks^9^ and 6 months; ^11^ and reliance on retrospectively reported pre-infection sleep data.^11^ More research is therefore needed to build our understanding of this relationship.

Understanding the role of sleep in long COVID risk and recovery is of key importance, both in preventing and treating long COVID, as well as preventing the known negative impacts of poor sleep, such as reduced quality of life^13^ and poorer health outcomes.^14^ With prospectively collected data on pre-infection sleep and regular long-term follow-up of post-infection sleep, the COVIDENCE UK cohort study (https://www.qmul.ac.uk/covidence) is well placed to examine the bidirectional relationship between sleep and long COVID. We therefore aimed to evaluate how pre-infection sleep quality is associated with risk of long COVID, and to track changes in sleep duration during the year after SARS-CoV-2 infection.

## Methods

### Study design and participants

COVIDENCE UK is a prospective, longitudinal, population-based observational study of COVID-19 in the UK population (https://www.qmul.ac.uk/covidence).^15^ Inclusion criteria were age 16 years or older and UK residence at enrolment, with no exclusion criteria. Participants were invited via a national media campaign via print and online newspapers, radio, television, social media, and online advertising. Enrolled participants completed an online baseline questionnaire and monthly follow-up questionnaires to capture information on potential symptoms of COVID-19, results of nose or throat swab tests for SARS-CoV-2, long COVID, and sleep characteristics. The study was launched on May 1, 2020, and closed to enrolment on Oct 6, 2021. The final COVIDENCE UK cohort was majority female (70.2%) and White (93.7%), with under-representation of people younger than 50 years, men, and minoritised ethnicities.^15^ This study is registered with ClinicalTrials.gov, NCT04330599. COVIDENCE UK was approved by Leicester South Research Ethics Committee (ref 20/EM/0117). All participants gave informed consent to take part in the study before enrolment.

To focus on community cases, we excluded participants who had been hospitalised with COVID-19. We additionally excluded participants with less than 12 weeks of follow-up after their infection date.

### SARS-CoV-2 infection

We defined evidence of a SARS-CoV-2 infection as a positive SARS-CoV-2 swab test or antibody test. As many people with long COVID were infected before widespread testing was available, we also included symptom-defined COVID-19 (based on the algorithm described by Menni et al^16^) with no concurrent negative SARS-CoV-2 test. Participants additionally reported the severity of their symptoms as mild (able to do most of usual activities), moderate (unable to do usual activities, but without requiring bedrest), or severe (requiring bedrest).

Infection dates were defined as the date of the test, for swab tests, and the date of symptom onset, for participants with symptom-defined COVID-19. Participants reporting positive antibody tests were asked to recall a suspected infection date (based on symptoms, swab test results, or close proximity to a COVID-19 case); participants unable to provide a date, who nonetheless reported probable COVID-19 symptoms^16^ before their antibody test, were assigned the symptom onset date as their infection date.

To enable inclusion of pre-infection data, time since infection was included as a categorical variable with the following categories: no infection, infection within the past month, infection 1–3 months before, infection 3–6 months before, infection 6–9 months before, and infection 9–12 months before. We chose these categories to enable a clearer distinction between the effects of the acute infection (≤1 month), ongoing symptomatic COVID-19 (1–3 months), and different phases of post-COVID-19 syndrome (>3 months).

We classified the likely SARS-CoV-2 variant as pre-Omicron or post-Omicron according to infection date, with the threshold set at Dec 20, 2021. ^17^

### Long COVID

Long COVID is variably defined by public health bodies as symptoms occurring more than 4 weeks after infection^18,19^ or more than 12 weeks after infection;^20^ there is still no international consensus. We therefore defined long COVID as at least two consecutive positive responses to the question “Would YOU say that you currently have ‘long COVID’, i.e. ongoing symptoms more than 4 weeks after the onset of proven or suspected COVID-19?” occurring at least 4 weeks after the participant’s first recorded infection date, and with at least one positive response more than 12 weeks after their first reported infection.

We constructed this definition to reflect a minimum frequency and persistence of ongoing symptoms—ie, symptoms occurring at least 4 weeks after the acute infection, with a duration of at least 2 months, and without having resolved by 12 weeks post infection. Participants were defined as not having long COVID if they never reported possible or suspected long COVID during follow-up.

### Sleep quality

We included measures of sleep that reflected timing, duration, efficiency, and variability, using a subset of questions from the Pittsburgh Sleep Quality Index (PSQI) designed to measure sleep duration and sleep efficiency (appendix p 2).^21^ The questions used in COVIDENCE UK focus on sleep duration and sleep efficiency. In each monthly questionnaire, participants are asked to report the average hours of actual sleep from the past month. Up to May, 2021, participants were also asked to report average sleep times (ie, when they turned out the lights in order to sleep) and rising times (ie, when they got up). Between October, 2020, and May, 2021, all participants were asked to additionally report whether they were experiencing problems sleeping, regardless of their infection status; after May, 2021, only participants reporting long COVID were asked to report on sleep problems.

We calculated the midpoint of sleep as the rising time minus half the hours spent asleep. We calculated sleep efficiency as the percentage of time in bed (ie, time between sleep times and rising times) spent asleep. Owing to strong collinearity between sleep duration and sleep efficiency, we constructed a sleep quality variable based on age-specific recommendations from the National Sleep Foundation (appendix figure S1). ^22,23^ For variability, we included the standard deviations of sleep midpoint, sleep duration, and sleep efficiency, calculated over all surveys completed at least 7 days before the infection date.

For pre-infection sleep, we used sleep data taken from each participant’s baseline survey, or their first pre-infection monthly survey if baseline measures were unavailable (<1% of participants).

### Covariates

We identified potential risk factors for long COVID from the literature (appendix table S1). Focusing on those that might be associated with sleep quality,^24,25^ we constructed directed acyclic graphs (DAGs) to identify potential confounders and to establish whether any adjustments could introduce bias (appendix figure S2). Based on our DAG, the minimally sufficient adjustment set comprised age; sex; body-mass index; socioeconomic status, as captured by the Index of Multiple Deprivation^26^ (in quartiles) and highest level of educational achievement (categorised as primary or secondary, higher or further, college or university, or postgraduate); baseline health, comorbidities, and anxiety or depression; and level of physical activity (categorised as 0 h, 1–3 h, or ≥4 h of vigorous physical activity per week). To increase statistical power, we used the number of comorbidities each participant had from the following list: diabetes, heart disease (coronary artery disease or heart failure), hypertension, respiratory disease (asthma or COPD), and immunosuppressants or organ transplantation. These comorbidities are present in the COVIDENCE UK dataset and have all been found to be associated with long COVID (appendix table S1). We adjusted separately for self-reported general health, and for presence of anxiety and depression, based on the Patient Health Questionnaire-4.^27^

For our analysis of post-infection sleep, we additionally included current employment status (defined as employed/working, not employed/working [unemployed, furloughed, or retired], or other) and vaccination status (any SARS-CoV-2 vaccines *vs* none).

### Statistical analysis

For our analysis of long COVID risk, we included all participants with evidence of a SARS-CoV-2 infection and pre-infection sleep data for all characteristics investigated. We carried out binary logistic regressions among infected participants, with pre-infection sleep characteristics as the exposures and long COVID as the outcome. Based on our DAG (appendix figure S2), we fitted an initial model without infection variables to estimate the total effect of pre-infection sleep characteristics on long COVID risk, and then included infection-related variables to explore how much infection severity mediated this effect, comparing model fit using the likelihood ratio test. We used orthogonal polynomial contrasts to test for linear trends in the ordinal sleep quality variable.

For our analysis of post-infection sleep, we included all participants with evidence of a SARS-CoV-2 infection who had data on pre-infection sleep quality and sleep problems. We carried out a repeated measures analysis of average monthly sleep duration, with time since infection as the exposure, using multilevel linear mixed models with random intercepts for participants. We censored cases at 1-year post infection or if they were reinfected. To explore how reporting long COVID manifests in sleep duration, we defined a three-level, time-varying, long COVID variable: does not report long COVID, reports long COVID without sleep problems, and reports long COVID with sleep problems. Further details on the model are included in the appendix (p 8).

We did several sensitivity analyses. First, to reduce the risk of misclassification, we restricted all analyses to people with test-confirmed COVID-19. Second, to explore the effect of sleep on different severities of long COVID, we calculated the total number of long COVID symptoms reported by each individual with long COVID, and restricted the long COVID risk analysis to those who reported more than five of the 16 symptoms included (ie, excluding the lowest quartile). Third, to ensure better comparability between baseline sleep data, we restricted the post-infection sleep analysis to participants with pre-infection sleep data from the first month that both sleep quality and sleep problems data were collected (October, 2020).

A previous COVIDENCE UK study has found that people who report a large number of symptoms consistent with long COVID do not necessarily report themselves as having long COVID, suggesting potential under-reporting of the condition. ^28^ We therefore carried out an exploratory analysis in which we extended the definition of long COVID to include participants who were less sure about their long COVID status, by counting the response “Don’t know / not sure” as a positive response. Finally, to assess the individual contributions of sleep duration and sleep efficiency towards long COVID risk, we carried out exploratory analyses substituting each variable in the model in the place of sleep quality.

We handled missing data with listwise deletion, under the assumption that data were missing at random, and that missingness was independent of the outcome variable, conditional on the covariates (appendix pp 21–24).^29^ We then explored potential selection bias using inverse probability weighting for logistic regressions.^29^ All data were analysed with Stata/MP 18. A p value of less than 0.05 was considered significant.

## Results

Between May, 2020, and March, 2021, we enrolled 3994 participants who were included in our analysis of long COVID risk (appendix figure S4). SARS-CoV-2 infections were recorded between July, 2020, and March, 2023. Participants were followed up for a median of 13.1 months (IQR 9.0–16.4) after their SARS-CoV-2 infection, during which 327 (8.2%) reported long COVID a median of five times (IQR 3–9; table 1). Compared with participants who did not report long COVID, those reporting long COVID were more likely to be younger and female, with lower levels of educational attainment, lower levels of physical activity, worse general health, higher BMI, and a greater burden of comorbidities (table 1). Participants reporting long COVID were also more likely to have been infected before the Omicron variant was dominant, and to have experienced a more severe acute infection, and were less likely to have been vaccinated at the time of infection (table 1).

**Table 1:** Baseline characteristics of participants included in the long COVID risk analysis.

|  | Reports long COVID<br>(n=327) | Does not report long COVID<br>(n=3667) |
| --- | --- | --- |
| **Sociodemographic and behavioural** |  |  |
| Age, years | 59.0 (49.1–66.7) | 63.4 (55.4–69.3) |
| Sex |  |  |
| Female | 252 (77.1%) | 2592 (70.7%) |
| Male | 75 (22.9%) | 1075 (29.3%) |
| Ethnicity |  |  |
| White | 312 (95.4%) | 3541 (96.6%) |
| Black, African, Caribbean, or Black British | 3 (0.9%) | 17 (0.5%) |
| South Asian | 4 (1.2%) | 47 (1.3%) |
| Mixed, multiple, or other | 8 (2.4%) | 62 (1.7%) |
| IMD decile | 7 (5–9) | 7 (5–9) |
| Highest educational level attained |  |  |
| Primary or secondary | 48 (14.7%) | 331 (9.0%) |
| Higher or further (A levels) | 53 (16.2%) | 484 (13.2%) |
| College or university | 131 (40.1%) | 1658 (45.2%) |
| Post-graduate | 95 (29.1%) | 1194 (32.6%) |
| Weekly hours of vigorous physical exercise |  |  |
| 0 | 144 (44.0%) | 1111 (30.3%) |
| 1–3 | 121 (37.0%) | 1689 (46.1%) |
| ≥4 | 62 (19.0%) | 867 (23.6%) |
| **Pre-infection sleep characteristics** |  |  |
| Sleep duration, h | 6.7 (1.2) | 7.0 (1.0) |
| Sleep efficiency | 80% (74–88) | 84% (76–90) |
| Sleep quality |  |  |
| Good | 96 (29.4%) | 1453 (39.6%) |
| Medium | 133 (40.7%) | 1459 (39.8%) |
| Medium–low | 78 (23.9%) | 669 (18.2%) |
| Low | 20 (6.1%) | 86 (2.3%) |
| Medications |  |  |
| Benzodiazepines | 4 (1.2%) | 11 (0.3%) |
| Non-benzodiazepine hypnotics and sedatives | 6 (1.8%) | 39 (1.1%) |
| **Clinical characteristics** |  |  |
| BMI, kg/m<sup>2</sup> |  |  |
| <25 | 126 (38.5%) | 2001 (54.6%) |
| 25 to <30 | 116 (35.5%) | 1129 (30.8%) |
| ≥30 | 85 (26.0%) | 537 (14.6%) |
| General health |  |  |
| Excellent | 48 (14.7%) | 1001 (27.3%) |
| Very good | 115 (35.2%) | 1662 (45.3%) |
| Good | 102 (31.2%) | 785 (21.4%) |
| Fair | 50 (15.3%) | 181 (4.9%) |
| Poor | 12 (3.7%) | 38 (1.0%) |
| Asthma or COPD | 69 (21.1%) | 516 (14.1%) |
| Autoimmune disease | 42 (12.8%) | 281 (7.7%) |
| Diabetes |  |  |
| Pre-diabetes | 13 (4.0%) | 101 (2.8%) |
| Diabetes | 23 (7.0%) | 141 (3.8%) |
| Heart disease | 15 (4.6%) | 101 (2.8%) |
| Hypertension | 72 (22.0%) | 755 (20.6%) |
| Immunosuppression or organ transplantation | 25 (7.6%) | 127 (3.5%) |
| PHQ-4 grade |  |  |
| Normal | 249 (76.4%) | 3273 (89.3%) |
| Mild | 61 (18.7%) | 311 (8.5%) |
| Moderate | 14 (4.3%) | 53 (1.4%) |
| Severe | 2 (0.6%) | 27 (0.7%) |
| **Acute infection and vaccination status** |  |  |
| Infection data type |  |  |
| Swab test | 312 (95.4%) | 3429 (93.5%) |
| Antibody test | 12 (3.7%) | 48 (1.3%) |
| Symptoms only | 3 (0.9%) | 190 (5.2%) |
| Likely SARS-CoV-2 variant |  |  |
| Pre-Omicron | 172 (52.6%) | 575 (15.7%) |
| Omicron | 155 (47.4%) | 3092 (84.3%) |
| Self-reported infection severity |  |  |
| Asymptomatic | 22 (6.7%) | 258 (7.0%) |
| Mildly unwell | 63 (19.3%) | 1402 (38.2%) |
| Moderately unwell | 99 (30.3%) | 1215 (33.1%) |
| Very unwell | 143 (43.7%) | 792 (21.6%) |
| Number of acute symptoms | 4 (3–5) | 2 (1–3) |
| Vaccinated at time of infection | 229 (70.0%) | 3443 (93.9%) |
| Follow-up since infection, months | 12.2 (8.5–15.3) | 16 (11.3–22.4) |

Data are n (%), mean(SD), or median (IQR). BMI=body-mass index. COPD=chronic obstructive pulmonary disease. IMD=Index of Multiple Deprivation. PHQ-4=Patient Health Questionnaire-4.

We found an association between pre-infection sleep quality and long COVID, with p-for-trend analyses suggesting higher risk of long COVID with lower sleep quality, regardless of infection severity (table 2). We additionally observed an association between greater variability in sleep efficiency and increased risk of long COVID, which was significant when adjusted for infection severity (table 2). Our results were similar after applying inverse probability weighting to correct for potential selection bias (data not shown). When considering sleep duration and sleep efficiency separately, we found lower sleep efficiency to be associated with higher risk of long COVID, but we lacked power to detect any association between sleep duration and long COVID (appendix table S4). Restricting our analysis to participants with test-confirmed COVID-19 did not substantially affect our results (table 2). Restricting our analysis to participants who may have a more severe form of long COVID (ie, reporting a greater number of long COVID symptoms) strengthened the association observed for both low-quality sleep and sleep efficiency variability (table 2). Extending our definition of long COVID to include participants with less certainty about their long COVID status attenuated some of the associations observed (table 2).

**Table 2:** Adjusted odds ratios for association between pre-infection sleep quality and risk of developing long COVID.

|  | Main analysis |  | Main analysis, adjusted for infection-related variables |  | Sensitivity 1: Test-positive only |  | Sensitivity 2: Higher symptom burden |  | Exploratory: Uncertain long COVID status |  |
| --- | --- | --- | --- | --- | --- | --- | --- | --- | --- | --- |
|  | OR (95% CI) | p value | OR (95% CI) | p value | OR (95% CI) | p value | OR (95% CI) | p value | OR (95% CI) | p value |
| Sleep quality |  |  |  |  |  |  |  |  |  |  |
| Good | 1 (ref) | .. | 1 (ref) | .. | 1 (ref) | .. | 1 (ref) | .. | 1 (ref) | .. |
| Medium | 1.37 (1.04–1.81) | 0.026 | 1.32 (0.98–1.78) | 0.072 | 1.27 (0.94–1.73) | 0.123 | 1.37 (0.95–1.97) | 0.088 | 1.29 (1.00–1.66) | 0.048 |
| Medium–low | 1.55 (1.12–2.16) | 0.009 | 1.49 (1.04–2.13) | 0.028 | 1.48 (1.03–2.13) | 0.036 | 1.93 (1.29–2.90) | 0.001 | 1.44 (1.07–1.95) | 0.018 |
| Low | 1.94 (1.11–3.38) | 0.020 | 2.02 (1.07–3.80) | 0.030 | 2.00 (1.05–3.81) | 0.035 | 2.58 (1.32–5.04) | 0.006 | 1.64 (0.92–2.91) | 0.093 |
| *p for linear trend* | .. |  | .. |  | .. | 0.026 | .. | 0.002 | .. | 0.076 |
| Sleep midpoint | 0.96 (0.87–1.07) | 0.476 | 1.00 (0.89–1.11) | 0.961 | 0.98 (0.87–1.10) | 0.698 | 0.99 (0.87–1.12) | 0.830 | 1.04 (0.95–1.12) | 0.043 |
| SD of mid-sleep point (10 min) | 1.00 (0.97–1.02) | 0.766 | 0.99 (0.96–1.02) | 0.432 | 0.99 (0.96–1.03) | 0.643 | 0.99 (0.96–1.02) | 0.470 | 0.99 (0.97–1.02) | 0.823 |
| SD of sleep efficiency (%) | 1.04 (1.00–1.09) | 0.077 | 1.07 (1.02–1.12) | 0.010 | 1.07 (1.02–1.12) | 0.010 | 1.07 (1.02–1.13) | 0.011 | 1.05 (1.01–1.10) | 0.926 |
| SD of time asleep (10 min) | 1.01 (0.93–1.09) | 0.797 | 0.98 (0.90–1.07) | 0.691 | 0.98 (0.90–1.07) | 0.683 | 1.02 (0.93–1.12) | 0.717 | 0.99 (0.92–1.07) | 0.128 |

The main analysis was adjusted for age, sex, body-mass index, socioeconomic status, general health, number of comorbidities, anxiety or depression, and physical activity. Adjustment for infection-related variables (infection severity, vaccination status, and variant) significantly improved model fit ( $p < 0.001$ ). Sensitivity and exploratory analyses are adjusted for infection-related variables. Sensitivity analysis 1 was restricted to participants with test-confirmed SARS-CoV-2 infection (3477 who did not report long COVID and 324 who did). Sensitivity analysis 2 restricted participants with long COVID to the 244 participants with a higher symptom burden (reporting at least six of the 16 symptoms investigated). The exploratory analysis extended the definition of long COVID to include the response “Don’t know / not sure” as a positive response, resulting in 483 participants reporting long COVID. Unadjusted ORs, and regression estimates for all covariates in the adjusted models, are presented in the appendix (tables S2–S4). OR=odds ratio. SD=standard deviation

Between May, 2020, and June, 2021, we enrolled 6860 participants who were included in our analysis of post-infection sleep (table 3). Their reported SARS-CoV-2 infections occurred between October, 2020, and March, 2023. Participants were followed up from enrolment for a median of 29 months (24–32). 2247 (32.9%) participants reported sleep problems at baseline (table 3).

**Table 3:** Baseline characteristics of participants included in the post-infection sleep analysis.

|  | Participants (n=6860) |
| --- | --- |
| **Sociodemographic and behavioural** |  |
| Age, years | 63.0 (54.2–69.1) |
| Sex |  |
| Female | 4898 (71.4%) |
| Male | 1962 (28.6%) |
| Ethnicity |  |
| White | 6595 (96.1%) |
| Black, African, Caribbean, or Black British | 28 (0.4%) |
| South Asian | 100 (1.5%) |
| Mixed, multiple, or other | 137 (2.0%) |
| IMD decile | 7 (5–9) |
| Highest educational level attained |  |
| Primary or secondary | 671 (9.8%) |
| Higher or further (A levels) | 956 (13.9%) |
| College or university | 3093 (45.1%) |
| Post-graduate | 2140 (31.2%) |
| Follow-up, months | 29.4 (24.4–32.5) |
| **Baseline sleep characteristics** |  |
| Sleep duration, h | 6.8 (1.0) |
| Sleep quality |  |
| Good | 2290 (33.4%) |
| Medium | 2731 (39.8%) |
| Medium–low | 1508 (22.0%) |
| Low | 331 (4.8%) |
| Reported sleep problems | 2247 (32.8%) |
| **Clinical characteristics** |  |
| BMI, kg/m<sup>2</sup> |  |
| <25 | 3416 (49.8%) |
| 25 to <30 | 2208 (32.2%) |
| ≥30 | 1236 (18.0%) |
| General health |  |
| Excellent | 1446 (21.1%) |
| Very good | 2855 (41.6%) |
| Good | 1794 (26.2%) |
| Fair | 631 (9.2%) |
| Poor | 134 (2.0%) |
| Asthma or COPD | 1156 (16.9%) |
| Autoimmune disease | 611 (8.9%) |
| Diabetes |  |
| Pre-diabetes | 6348 (92.5%) |
| Diabetes | 212 (3.1%) |
| Heart disease | 259 (3.8%) |
| Hypertension | 1502 (21.9%) |
| Immunosuppression or organ transplantation | 322 (4.7%) |
| Medications |  |
| Benzodiazepines | 29 (0.4%) |
| Non-benzodiazepine hypnotics and sedatives | 79 (1.2%) |
| **Acute infection and long COVID** |  |
| Infection data type |  |
| Swab test | 6353/6855 (92.7%) |
| Antibody test | 123/6855 (1.8%) |
| Symptoms only | 379/6855 (5.5%) |
| Self-reported infection severity |  |
| Asymptomatic | 445 (6.5%) |
| Mildly unwell | 2171 (31.6%) |
| Moderately unwell | 2244 (32.7%) |
| Very unwell or hospitalised | 2000 (29.2%) |
| Ever reports long COVID | 333 (4.9%) |

Data are n (%), mean (SD), or median (IQR). BMI=body-mass index. IMD=Index of Multiple Deprivation. PHQ-4=Patient Health Questionnaire-4.

We found that sleep duration increased slightly in the month immediately following a SARS-CoV-2 infection (figure; appendix tables S5, S6). The size of this increase was dependent on the severity of the acute infection, with no increase for people who were asymptomatic or had mild disease (figure; appendix tables S5, S6). Across all infection severities, no significant changes in sleep duration were observed more than 1 month after infection (figure; appendix tables S5, S6). Greater fluctuations in sleep duration were seen when assessing participants according to long COVID status. Participants who reported long COVID without sleep problems experienced a greater increase in sleep duration in the first month after infection, and a slower return to pre-infection sleep duration (figure; appendix tables S5, S6). Those who reported long COVID with sleep problems experienced a slight decrease in sleep duration between 1 and 6 months after infection, after which sleep duration returned to pre-infection levels (figure; appendix tables S5, S6). However, the number of people long COVID with sleep problems were small, leading to greater uncertainty in our estimates.

When restricting our analyses to participants with pre-infection sleep data from October, 2020, we saw a greater reduction and more extended fluctuations in sleep duration among people reporting long COVID with sleep problems (appendix table S6). No other substantial changes were observed in our results across our sensitivity analyses (appendix tables S6).

## Discussion

In this large population-based prospective study, we found that poor-quality sleep—defined by the combination of sleep duration and sleep efficiency—and greater variability over time in sleep efficiency before SARS-CoV-2 infection were both associated with increased risk of reporting long COVID. By contrast, we found no evidence that pre-infection sleep timing nor variability in sleep duration affect risk of long COVID. We observed an increase in sleep duration over the month following a SARS-CoV-2 infection, with greater increases among participants with more severe acute infections. After 1 month, however, sleep duration largely returned to pre-infection levels. Participants reporting long COVID showed greater fluctuations and slower recovery of pre-infection sleep duration, with the direction of change depending on whether their long COVID presented with sleep problems. Estimated changes to sleep were all less than 25 minutes per night and did not last more than 6 months after infection.

Short sleep duration and sleep disturbances have been found to be associated with increased incidence of upper respiratory tract infections, including COVID-19,^8,30,31^ as well as poorer outcomes from these infections.^8,31^ Our longitudinal analysis extends these findings to show the bidirectional nature of the relationship between sleep and COVID-19 within the same cohort. A large cross-sectional study^32^ of both community and hospitalised COVID-19 cases has previously shown a bidirectional relationship between insomnia and long COVID, reporting increased risk of long COVID among people with pre-pandemic insomnia and a higher prevalence of post-infection insomnia among people with long COVID. However, this study relied on retrospectively reported sleep data, and did not benefit from repeated measures of sleep after infection to track whether insomnia symptoms resolved over time.

We show that the quality of pre-infection sleep predicts long COVID risk, independently of the severity of the acute infection, supporting and extending findings from a previous study^9^ done exclusively in women. While the mechanisms behind long COVID remain unknown, sleep quality and disturbances are known to play a part in many of the candidate mechanisms under investigation, ^1^ such as persistent inflammation, ^33^ changes to the gut microbiota, ^34^ and autoimmunity. ^33^ Our sleep quality measure combined the effects of both sleep efficiency and sleep duration, but exploratory analyses suggested that low and inconsistent sleep efficiency—which can reflect a range of sleep disturbances, such as increased sleep latency and fragmented sleep—was driving the relationship observed. This is in line with other studies, which have found increased risk of long COVID associated with pre-infection sleep disturbances, ^9–11^ whereas results on sleep duration are inconsistent^9,10^ and do not always consider the simultaneous effect of sleep efficiency. ^35^ Together, these findings suggest short sleep duration, on its own, is unlikely to substantially increase the risk of long COVID; instead, the focus should be on ensuring consistently good-quality sleep to aid recovery from future infections.

The small increase in sleep duration we observed in the month after SARS-CoV-2 infection, followed by a return to pre-infection sleep duration, is consistent with recovery from acute respiratory infection,^36^ and will reflect the experience of the many people who recover well from SARS-CoV-2 infection. However, our finding that participants with and without long COVID recovered their pre-infection sleep duration within 6 months after infection contrasts with the few other studies that have looked at sleep duration specifically. A cross-sectional survey in both hospitalised and non-hospitalised participants showed a decrease in satisfaction with sleep duration 6 months after infection;^37^ however, this subjective measure of pre-infection sleep was retrospectively reported and the duration itself was not measured. Wearable devices have been used to objectively record sleep duration, showing different results in community and hospitalised cohorts. At 6 months after infection, a cohort of mostly non-hospitalised participants had a mean sleep duration that was 17 minutes shorter than that of uninfected controls,^38^ whereas hospitalised participants from the PHOSP cohort reported sleeping for an hour longer at 7 months after infection than a cohort of non-hospitalised controls.^3^ Our findings may reflect a quicker recovery among less severe cases of COVID-19, as well as the benefit of comparing post-infection sleep duration with prospectively collected pre-infection sleep data; indeed, another study in the PHOSP cohort showed no difference in sleep duration at 8 months after discharge when compared with a different control cohort.^39^

Although all groups in our analysis recovered their pre-infection sleep duration within 6 months after infection, some participants continued to report long COVID with sleep problems after this period. There is substantial evidence on sleep disturbances after COVID-19,^2,40^ with an estimated prevalence of 25% after 12 months,^2^ increasing to 50% among people hospitalised with COVID-19.^41^ Some facets of sleep health may therefore continue to be affected once pre-infection sleep duration has been recuperated, and further research is needed to better understand how longer-term sleep problems manifest in long COVID.

This population-based study has several strengths. Our use of prospectively collected, pre-infection sleep data strengthens our estimates of pre-infection sleep; most studies to date have relied on retrospectively reported pre-infection sleep data,^3,32,35,37,38^ and research has shown that people asked to retrospectively recall their sleep before the pandemic estimated that they slept much better than they actually did.^6^ Our extended follow-up enabled us to capture long COVID symptoms presenting at any point after infection, and to record sleep duration over the entire year after infection, in contrast with other studies that have relied on a cross-sectional approach,^32,37^ infrequent assessments,^10^ or shorter follow-up.^38^ Monthly responses from our participants meant that we were able to track changes in sleep duration with detail that would have been missed if observations had been further apart, while simultaneously allowing us to adjust for time-varying factors that may affect people’s sleep habits—particularly in the context of a pandemic that limited people’s movement and employment.

This study also has various limitations. First, we relied on self-reported measures of sleep, rather than actigraphy or polysomnography data, meaning our response data were subjective and subject to recall bias. Additionally, as sleep data were reported as monthly averages, we will have missed smaller fluctuations. Second, we were unable to assess changes to sleep quality—a variable defined by both sleep duration and sleep efficiency—after SARS-CoV-2 infection; this is because questions used to calculate sleep efficiency were removed from the COVIDENCE UK questionnaire in May, 2021, to reduce the burden on participants. Sleep is a multidimensional entity, and the recovery we observed in sleep duration may not necessarily be reflected in other dimensions of sleep, which should be evaluated in future studies. Third, our study is restricted to community cases of COVID-19, meaning our findings may not reflect the experiences of people with severe disease. However, several studies in hospitalised people have been published;^5,39^ as long COVID occurs across all severities of the initial disease,^1^ it is important to assess its impact in the community as well as in those hospitalised. Fourth, as the severity of the acute infection may also be associated with pre-infection sleep,^8^ including people with symptomatic COVID-19 in our reference group may have underestimated the impact of pre-infection sleep on long COVID risk. However, we had too few asymptomatic participants to exclude symptomatic participants from the reference group. Fifth, COVIDENCE UK is a self-selected cohort,^15^ and so certain groups—such as older age groups, women, and people of White ethnicity—are over-represented in our study, which may limit the generalisability of our results. Sixth, we analysed our data under the assumption that data were missing at random, which may have biased our results. However, sensitivity analyses using inverse probability weights to correct for potential selection bias resulted in similar findings. Finally, as with any observational study, we cannot rule out the possibility of unmeasured or residual confounding influencing our findings. However, the richness of the data collected in COVIDENCE UK means that we were able to control for various potential confounders in our analyses.

In conclusion, our study highlights the bidirectional relationship between sleep and long COVID, illustrating how poor-quality sleep before SARS-CoV-2 infection may increase the risk of long COVID, and describing changes to sleep duration after SARS-CoV-2 infection. The modest changes to sleep duration we observed in these non-hospitalised cases of COVID-19 largely resolved within 6 months, potentially reflecting a quicker recovery of pre-infection sleep characteristics among less severe infections.

## Contributors

GV, ARM, JB, and CJ conceived the analysis. GV and MT contributed to data management, and have directly accessed and verified the data. Statistical analyses were done by GV, with input from all authors. GV wrote the first draft of the report. All authors provided critical conceptual input, interpreted the data analysis, and read and approved the final draft. All authors had full access to all the data in the study and had final responsibility for the decision to submit the manuscript for publication.

## Data sharing

De-identified participant data will be made upon reasonable request to the corresponding author.

## Data Availability

De-identified participant data will be made upon reasonable request to the corresponding author.

## Acknowledgments

COVIDENCE UK has received support from Barts Charity (MGU0459, MGU0466), Pharma Nord, the Fischer Family Foundation, DSM Nutritional Products, the Epilepsy Foundation, the Karl R Pfleger Foundation, the AIM Foundation, Synergy Biologics, Cytoplan, the UK NIHR Clinical Research Network (52255; 52257), the Health Data Research UK BREATHE Hub, the UKRI Industrial Strategy Challenge Fund (MC_PC_19004), Thornton & Ross, Warburtons, Matthew Isaacs (personal donation), Barbara Boucher (personal donation), and Hyphens Pharma. MT was supported by Barts Charity (MGU0570). JB was supported by a Medical Research Council transition support fellowship (MR/T032529/1). CJ is funded by an Engineering and Physical Sciences Research Council (EPSRC) Mathematics Doctoral Training Partnership (EP/W523884/1). The views expressed are those of the authors and not necessarily those of the funders. We thank all participants of COVIDENCE UK, and the following organisations who supported study recruitment: Asthma UK/British Lung Foundation, the British Heart Foundation, the British Obesity Society, Cancer Research UK, Diabetes UK, Future Publishing, Kidney Care UK, Kidney Wales, Mumsnet, the National Kidney Federation, the National Rheumatoid Arthritis Society, the North West London Health Research Register (DISCOVER), Primary Immunodeficiency UK, the Race Equality Foundation, SWM Health, the Terence Higgins Trust, and Vasculitis UK. We also thank Sheena Maltby (QMUL, London, UK) for her support of both the COVIDENCE UK study and its participants.

---

**Figure:**
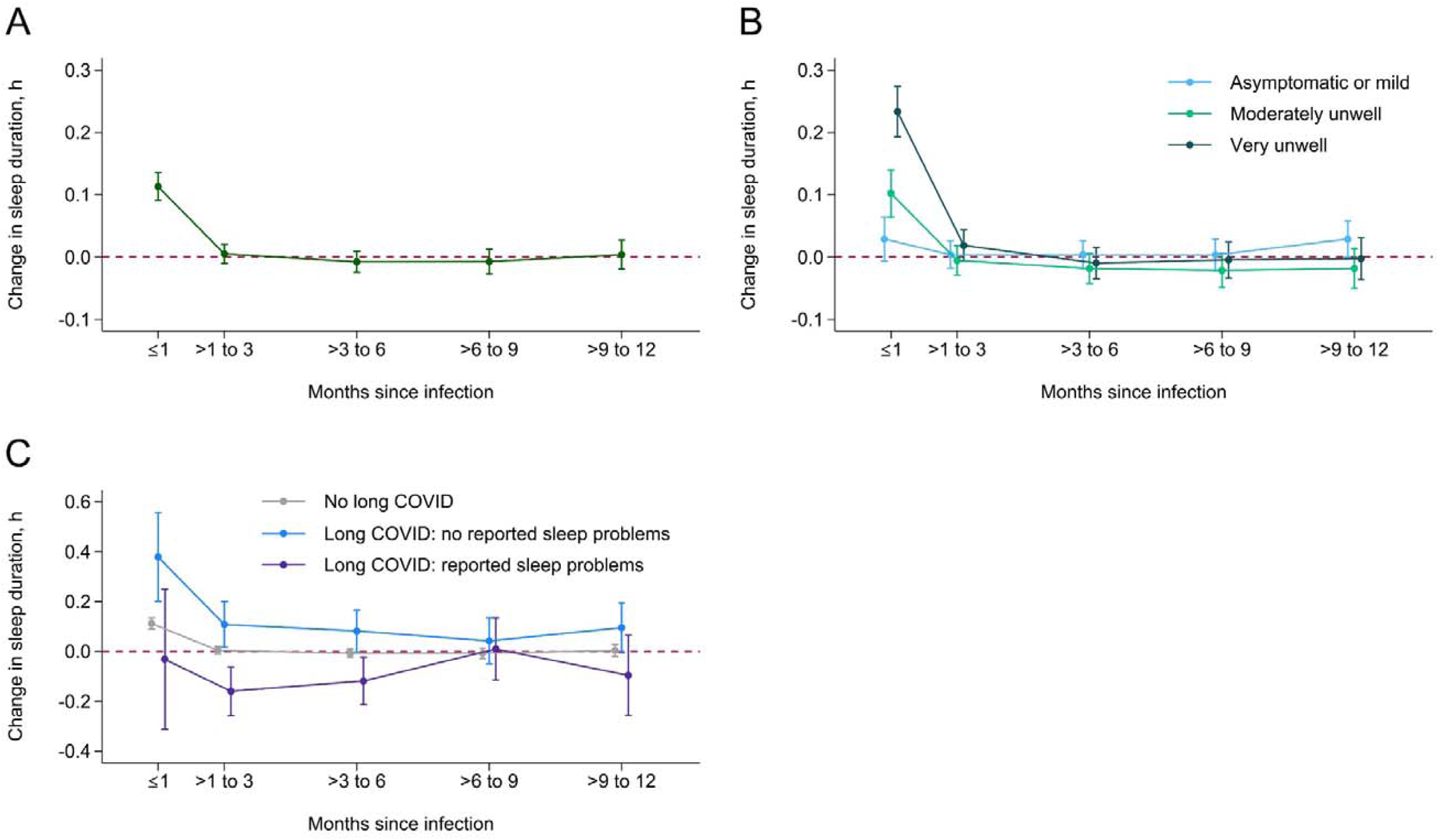
Changes in sleep duration after SARS-CoV-2 infection overall (A) and by infection severity (B) and long COVID status (C) Figure shows estimated changes to sleep duration over time in all participants (A), and according to severity of the acute infection (B) and long COVID status (C). Long COVID status is further categorised according to whether participants were reporting long COVID with sleep problems or long COVID without sleep problems. Estimates shown are contrasts of predictive margins from the regression models, with 95% CIs. Regression estimates are included in the appendix (tables S5, S6). Red dashed line indicates pre-infection sleep duration.

