## Appendix for "Bidirectional relationship between sleep problems and long COVID: a longitudinal analysis of data from the COVIDENCE UK study"

### **Pittsburgh Sleep Quality Index**

The Pittsburgh Sleep Quality Index (PSQI) assesses seven components of sleep: subjective sleep quality, sleep latency, sleep duration, habitual sleep efficiency, sleep disturbances, use of sleeping medication, and daytime dysfunction. Participants are ask to report their usual sleep habits for the past month.

The COVIDENCE UK questionnaire included questions 1, 3, and 4 from the PSQI, which are designed to assess sleep duration and sleep efficiency.

1. During the past month, what time have you usually gone to bed at night?

3. During the past month, what time have you usually gotten up in the morning?

4. During the past month, how many hours of actual sleep did you get at night? (This may be different than the number of hours you spent in bed.)

|  |  | Hours asleep | | | | | | | | | | | | | | | | | | | | | | | | | | | | | | |  | |  |
| --- | --- | --- | --- | --- | --- | --- | --- | --- | --- | --- | --- | --- | --- | --- | --- | --- | --- | --- | --- | --- | --- | --- | --- | --- | --- | --- | --- | --- | --- | --- | --- | --- | --- | --- | --- |
|  | Young adults | <5 | 5 | 6 | 7 | | 8 | | 9 | 10 | 11 | >11 | <5 | 5 | 6 | 7 | 8 | 9 | 10 | 11 | >11 | <5 | 5 | 6 | 7 | 8 | | 9 | 10 | 11 | >11 | | Adults  and older adults | |  |
| Sleep efficiency | ≤64% |  |  |  |  | |  | |  |  |  |  |  |  |  |  |  |  |  |  |  |  |  |  |  |  | |  |  |  |  | | ≤64% | | Sleep efficiency |
|  | 65–74% |  |  |  |  | |  | |  |  |  |  |  |  |  |  |  |  |  |  |  |  |  |  |  |  | |  |  |  |  | | 65–74% | |  |
|  | 75–84% |  |  |  |  | |  | |  |  |  |  |  |  |  |  |  |  |  |  |  |  |  |  |  |  | |  |  |  |  | | 75–84% | |  |
|  | ≥85% |  |  |  |  | |  | |  |  |  |  |  |  |  |  |  |  |  |  |  |  |  |  |  |  | |  |  |  |  | | ≥85% | |  |
|  |  | Young adult (≤25 years) | | | | | | | | | | | Adult (26–64 years) | | | | | | | | | Older adult (≥65 years) | | | | | | | | | | |  | |  |
|  |  | Sleep quality | | | | | |  | | | | |  | | | | | | | | |  | | | | | NSF category | | | | | | |  | |
|  |  | Good | | | |  | | Both sleep efficiency and hours asleep within 'recommended' levels | | | | | | | | | | | | | |  | | | | | Recommended | | | | |  | |  | |
|  |  | Medium | | | |  | | Either sleep efficiency or hours asleep within 'inconclusive' levels | | | | | | | | | | | | | |  | | | | | Inconclusive | | | | |  | |  | |
|  |  | Medium–low | | | |  | | Either sleep efficiency or hours asleep within 'not recommended' levels | | | | | | | | | | | | | |  | | | | | Not recommended | | | | |  | |  | |
|  |  | Low | | | |  | | Both sleep efficiency and hours asleep within 'not recommended' levels | | | | | | | | | | | | | |  | | | | | | | | | | |  | |  |

### ***Figure S1:* Sleep quality, categorised according to recommendations on sleep efficiency and duration from the NSF**

The grid shows which combinations of sleep efficiency and duration are included in each sleep quality category. The NSF categories for sleep efficiency and duration, by age, are shown around the edges of the grid. NSF=National Sleep Foundation.

### ***Table S1:* Potential long COVID risk factors**

| **Risk factors** | **Studies** |
| --- | --- |
| Sociodemographic and behavioural | |
| Age | ^5-15^ |
| Sex | ^5-10,12-27^ |
| Ethnicity | ^5,15,28^ |
| Educational attainment | ^29,30^ |
| Socioeconomic status or financial security | ^7,15,31,32^ |
| Smoking status | ^5,9,14,15,25,26^ |
| Vaping status | ^5^ |
| Pre-infection physical activity | ^33^ |
| Clinical characteristics | |
| BMI | ^5,15,18,23,33-36^ |
| Baseline general health / disability | ^6-8,13,19,26,37^ |
| Asthma | ^22^ |
| Diabetes | ^23,30,36,38^ |
| Heart disease | ^22^ |
| Hypertension | ^9,24,27,30^ |
| Organ transplant | ^36^ |
| Psychological conditions | ^13,32,39,40^ |
| Respiratory disease | ^15,33^ |
| Sleeping problems | ^41,42^ |
| Immunosuppressants | ^20^ |
| Vitamin D | ^43^ |
| Acute infection and vaccination status | |
| SARS-CoV-2 variant | ^32^ |
| COVID-19 severity | ^5,6,8,9,13,16,18,19,22,26,27,29,35-37,44^ |
| Number of symptoms during acute phase | ^11,32,37^ |
| Anosmia | ^10,11,47,48^ |
| Diarrhoea | ^10,29,49^ |
| Dyspnoea | ^10,29,35,47^ |
| Fatigue | ^11,47,48^ |
| Headache | ^35,47,48,50^ |
| Myalgia | ^9,11,29,35^ |
| Rash | ^10^ |
| Throat pain | ^29^ |
| Repeat SARS-CoV-2 infections | ^45,46^ |
| Vaccination status | ^10,26,37,44,51-53^ |

BMI=body-mass index.

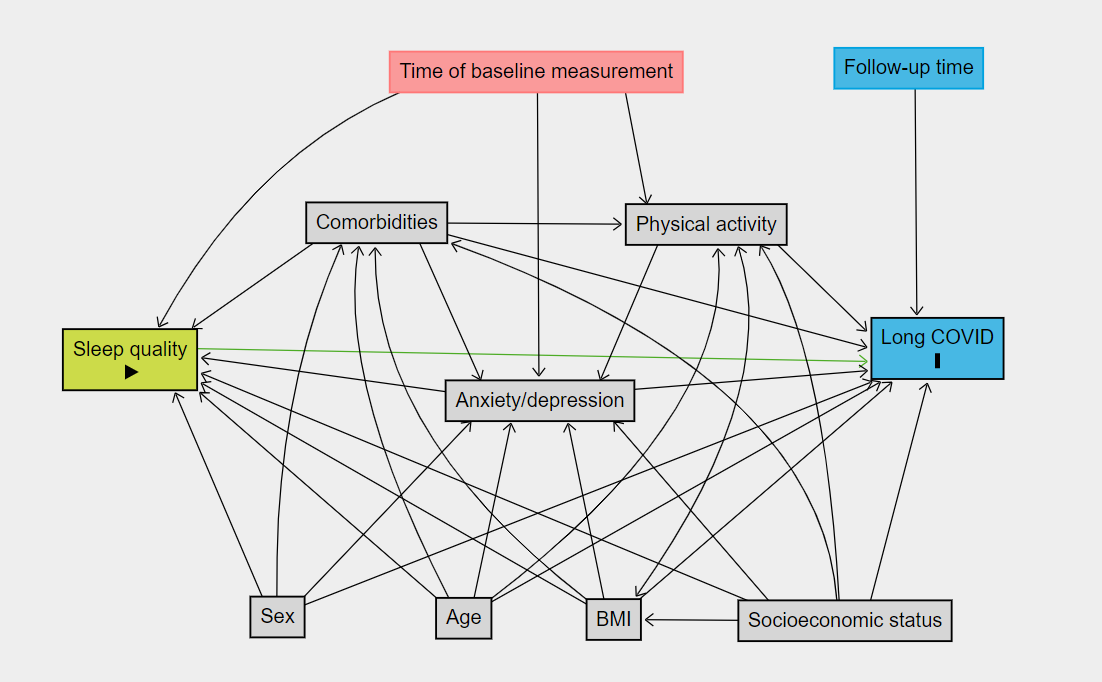

### ***Figure S2:* DAGs for long COVID risk**

**A**

**B**

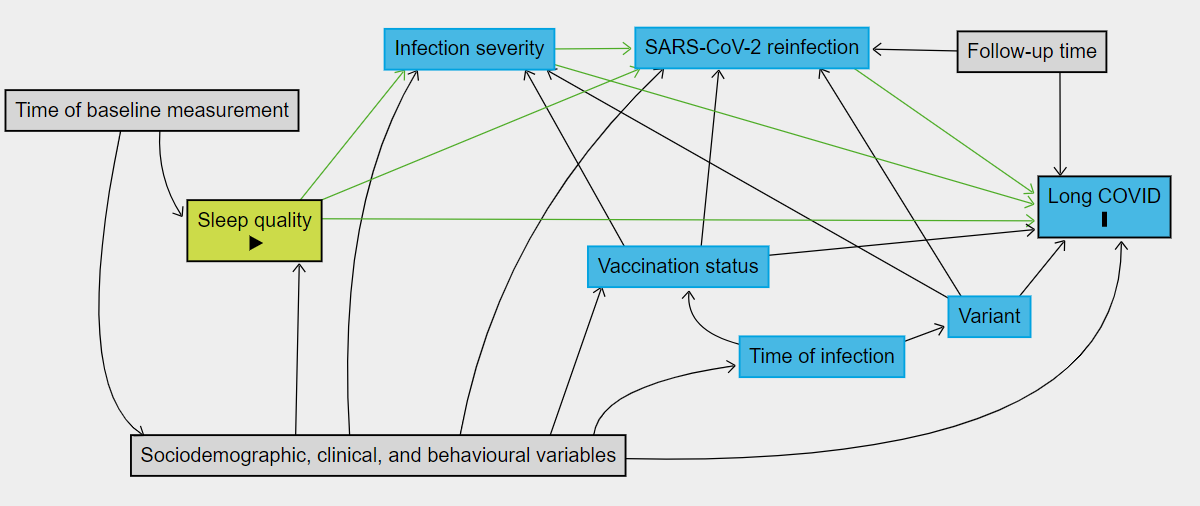

Directed acyclic graph (DAG) A shows the model including only sociodemographic, behavioural, and clinical variables. All variables except for the time variables need to be adjusted for to avoid confounding; adjustment for both time variables, however, does not introduce confounding. Infection-related variables are shown in DAG B, where we assume the socioeconomic and clinical variables in DAG A have been adjusted for. Under model assumptions (namely, that sleep quality affects infection severity through the immune response [unobserved]), adjustment for infection variables is not required to estimate the total effect of sleep quality on risk of long COVID.

To explore how much the effect of sleep quality is driven by subsequent infection severity, we can further adjust for infection variables, alongside vaccination status and variant to avoid introducing confounding.

To estimate the direct effect of sleep quality on long COVID risk, we would need to additionally adjust for SARS-CoV-2 reinfection. However, we were unable to do so successfully, as participants who do not report long COVID had a longer time period over which to report reinfections, which introduced bias into the model.

As DAGs are, by definition, directed and acyclic, we should not permit bidirectional causal relationships. However, in this diagram, we include a bidirectional relationship between BMI and physical activity, and BMI and anxiety and depression, to recognise that when these three variables are measured at the same timepoint, the causal relationships between the three are unclear. However, either direction of these relationships do not introduce confounding if all three variables are adjusted for.

Importantly, some relationships may be cyclical in nature when considered more broadly (eg, people experiencing depression may be less likely to do physical activity, which may in turn help to perpetuate depression). Here, we present the directions we believe are most likely to be present in our data, based on the context of the study.

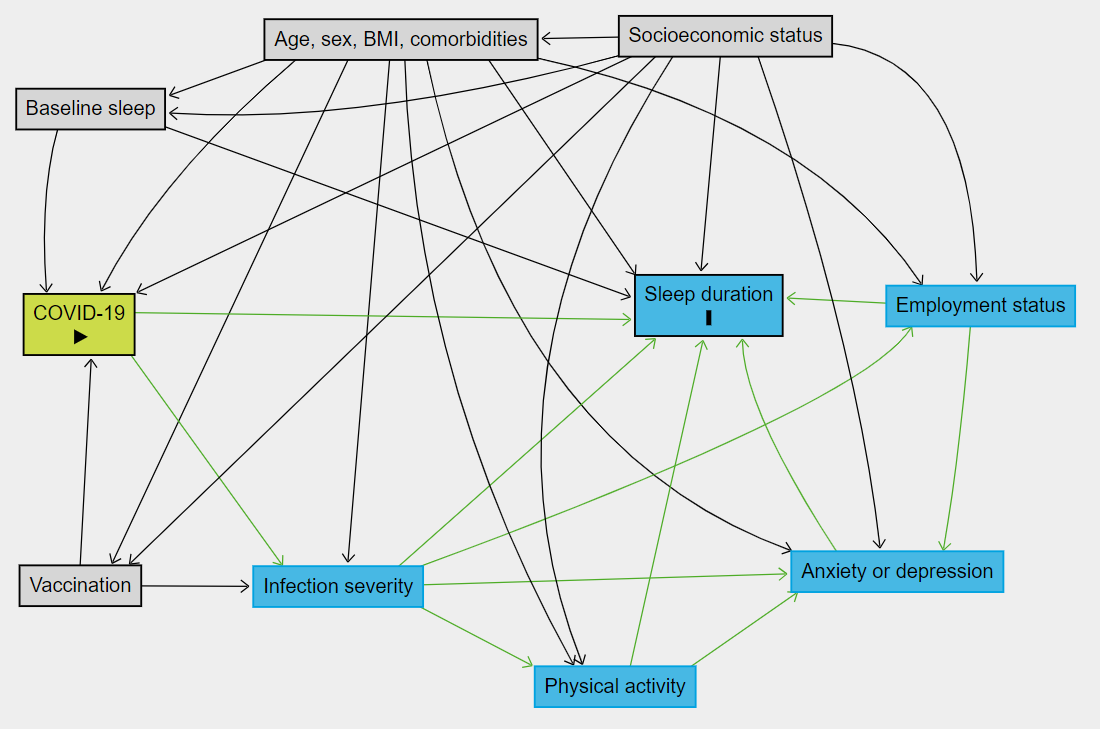

### ***Figure S3:* DAG for post-infection sleep**

The DAG shows necessary adjustments for baseline sleep, age, sex, BMI, comorbidities, socioeconomic status, vaccination status, and calendar time. Infection severity can only be explored in analyses without control participants. Time-varying variables such as employment status, anxiety or depression, and physical activity do not need to be adjusted for to explore the total effect of COVID-19 on sleep duration over time; however, the DAG suggests they can all be adjusted for without introducing bias as long as infection severity is also adjusted for.

### **Further details on multilevel linear mixed models**

We carried out a repeated measures analysis of average monthly sleep duration, with time since infection as the exposure, using multilevel linear mixed models with random intercepts for participants.

As we expected observations among each participant to be sequentially correlated, we used an autoregressive (AR) covariance structure. We used the Aikake information criterion (AIC) and the Bayesian information criterion (BIC) to compare models with different lags, choosing AR(2) for our final covariance structure. To account for the effect different moments during the pandemic may have had on factors such as sleep behaviours, anxiety or depression, and fatigue, we included crossed random effects between the questionnaire number (an indicator of month and year) and participant identifier. We included a random slope for time since infection, to account for individual variation on how much an infection may impact sleep.

To explore how reporting long COVID manifests in sleep duration, we defined a three-level, time-varying, long COVID variable: does not report long COVID, reports long COVID without sleep problems, and reports long COVID with sleep problems. We added interactions between time since infection and infection severity, as we hypothesised that more severe infections may have a longer-lasting effect on sleep. We also included an interaction between time since infection and long COVID status, as we hypothesised that the way long COVID manifests may change over time.

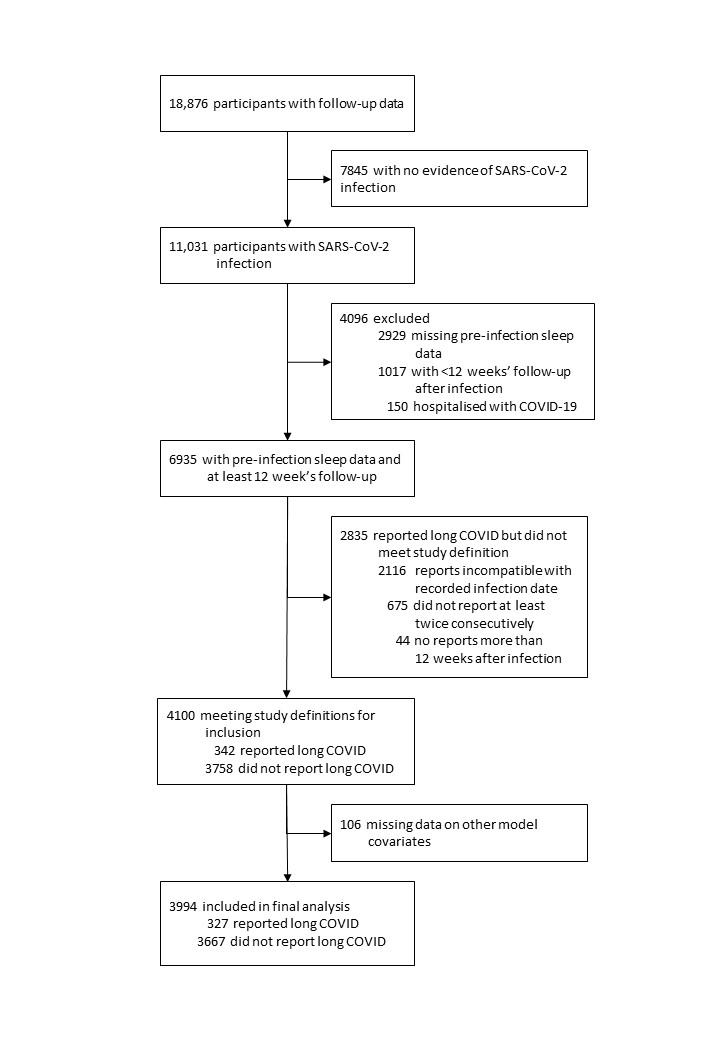

### ***Figure S4:* Participant flow diagram: analysis on long COVID risk**

### ***Table S2:* Regression estimates from logistic regression: unadjusted odds ratios and main analysis**

|  | **Unadjusted** | | **Main model** | |
| --- | --- | --- | --- | --- |
|  | OR (95% CI) | p value | OR (95% CI) | p value |
| Sleep quality |  |  |  |  |
| Good | 1 (ref) |  | 1 (ref) | ·· |
| Medium | 1.39 (1.07–1.82) | 0.015 | 1.32 (0.98–1.78) | 0.072 |
| Medium–low | 1.73 (1.27–2.35) | <0.001 | 1.49 (1.04–2.13) | 0.028 |
| Low | 3.72 (2.23–6.19) | <0.001 | 2.02 (1.07–3.80) | 0.030 |
| Sleep midpoint | 0.99 (0.90–1.10) | 0.914 | 1.00 (0.89–1.11) | 0.961 |
| SD of mid-sleep point (10 min) | 1.02 (1.00–1.04) | 0.037 | 0.99 (0.96–1.02) | 0.432 |
| SD of sleep efficiency (%) | 1.09 (1.06–1.12) | <0.001 | 1.07 (1.02–1.12) | 0.010 |
| SD of time asleep (10 min) | 1.14 (1.08–1.20) | <0.001 | 0.98 (0.90–1.07) | 0.691 |
| Age, years | ·· | ·· | 0.99 (0.98–1.00) | 0.016 |
| Sex |  |  |  |  |
| Female | ·· | ·· | 1.13 (0.84–1.54) | 0.420 |
| Male | ·· | ·· | 1 (ref) | ·· |
| Highest educational level attained |  |  |  |  |
| Primary or secondary | ·· | ·· | 1.71 (1.11–2.62) | 0.015 |
| Higher or further (A levels) | ·· | ·· | 1.41 (0.95–2.10) | 0.087 |
| College or university | ·· | ·· | 0.99 (0.73–1.34) | 0.964 |
| Post-graduate | ·· | ·· | 1 (ref) | ·· |
| Quartiles of IMD decile |  |  |  |  |
| Q4 (least deprived) | ·· | ·· | 1 (ref) | ·· |
| Q3 | ·· | ·· | 0.74 (0.52–1.03) | 0.078 |
| Q2 | ·· | ·· | 0.97 (0.69–1.37) | 0.874 |
| Q1 (most deprived) | ·· | ·· | 0.96 (0.68–1.36) | 0.822 |
| BMI (kg/m²) | ·· | ·· | 1.02 (0.99–1.04) | 0.130 |
| General health |  |  |  |  |
| Excellent | ·· | ·· | 1 (ref) | ·· |
| Very good | ·· | ·· | 1.19 (0.82–1.73) | 0.348 |
| Good | ·· | ·· | 1.64 (1.09–2.45) | 0.017 |
| Fair | ·· | ·· | 2.77 (1.65–4.64) | <0.001 |
| Poor | ·· | ·· | 1.84 (0.76–4.49) | 0.179 |
| Number of comorbidities | ·· | ·· | 1.21 (1.04–1.41) | 0.013 |
| Any depression or anxiety (PHQ-4) | ·· | ·· | 1.08 (0.75–1.57) | 0.671 |
| Vigorous physical exercise, h per week |  |  |  |  |
| 0 h | ·· | ·· | 1 (ref) | ·· |
| 1–3 h | ·· | ·· | 0.80 (0.60–1.07) | 0.131 |
| ≥4 h | ·· | ·· | 0.81 (0.57–1.16) | 0.251 |
| Self-reported infection severity |  |  |  |  |
| Asymptomatic | ·· | ·· | 1 (ref) | ·· |
| Mildly unwell | ·· | ·· | 1.16 (0.67–2.00) | 0.596 |
| Moderately unwell | ·· | ·· | 2.20 (1.29–3.75) | 0.004 |
| Very unwell | ·· | ·· | 4.17 (2.46–7.08) | <0.001 |
| Vaccinated at time of infection | ·· | ·· | 0.56 (0.38–0.82) | 0.003 |
| Variant |  |  |  |  |
| Pre-Omicron | ·· | ·· | 1 (ref) | ·· |
| Post-Omicron | ·· | ·· | 0.29 (0.20–0.41) | <0.001 |
| Baseline questionnaire | ·· | ·· | 1.04 (0.98–1.10) | 0.194 |
| Follow-up from SARS-CoV-2 infection, months | ·· | ·· | 1.04 (1.01–1.06) | 0.001 |

Unadjusted ORs were obtained from separate logistic regression models for each sleep variable. Main analysis shown is adjusted for BMI=body-mass index. IMD=Index of Multiple Deprivation. OR=odds ratio. PHQ-4=Patient Health Questionnaire-4.

### ***Table S3:* Regression estimates from logistic regression: sensitivity and exploratory analyses**

|  | **Sensitivity 1** | | **Sensitivity 2** | | **Exploratory** | |
| --- | --- | --- | --- | --- | --- | --- |
|  | OR (95% CI) | p value | OR (95% CI) | p value | OR (95% CI) | p value |
| Sleep quality |  |  |  |  |  |  |
| Good | 1 (ref) | ·· | 1 (ref) | ·· | 1 (ref) | ·· |
| Medium | 1.27 (0.94–1.73) | 0.036 | 1.37 (0.95–1.97) | 0.001 | 1.29 (1.00–1.66) | 0.048 |
| Medium–low | 1.48 (1.03–2.13) | 0.035 | 1.93 (1.29–2.90) | 0.006 | 1.44 (1.07–1.95) | 0.018 |
| Low | 2.00 (1.05–3.81) | 0.698 | 2.58 (1.32–5.04) | 0.830 | 1.64 (0.92–2.91) | 0.093 |
| Sleep midpoint | 0.98 (0.87–1.10) | 0.643 | 0.99 (0.87–1.12) | 0.470 | 1.04 (0.95–1.12) | 0.416 |
| SD of mid-sleep point (10 min) | 0.99 (0.96–1.03) | 0.010 | 0.99 (0.96–1.02) | 0.011 | 0.99 (0.97–1.02) | 0.627 |
| SD of sleep efficiency (%) | 1.07 (1.02–1.12) | 0.683 | 1.07 (1.02–1.13) | 0.717 | 1.05 (1.01–1.10) | 0.015 |
| SD of time asleep (10 min) | 0.98 (0.90–1.07) | 0.081 | 1.02 (0.93–1.12) | 0.020 | 0.99 (0.92–1.07) | 0.861 |
| Age, years | 0.99 (0.98–1.00) | <0.001 | 0.98 (0.97–1.00) | <0.001 | 0.99 (0.98–1.00) | 0.085 |
| Sex |  |  |  |  |  |  |
| Female | 1.11 (0.81–1.52) | 0.514 | 1.17 (0.81–1.68) | 0.400 | 1.13 (0.87–1.46) | 0.357 |
| Male | 1 (ref) | ·· | 1 (ref) | ·· | 1 (ref) | ·· |
| Highest educational level attained |  |  |  |  |  |  |
| Primary or secondary | 1.71 (1.10–2.65) | 0.016 | 2.25 (1.38–3.68) | 0.001 | 1.36 (0.93–2.00) | 0.114 |
| Higher or further (A levels) | 1.44 (0.96–2.16) | 0.075 | 1.64 (1.03–2.61) | 0.037 | 1.33 (0.94–1.87) | 0.104 |
| College or university | 1.00 (0.74–1.36) | 0.991 | 1.16 (0.81–1.66) | 0.418 | 1.10 (0.85–1.41) | 0.483 |
| Post-graduate | 1 (ref) | ·· | 1 (ref) | ·· | 1 (ref) | ·· |
| Quartiles of IMD decile |  |  |  |  |  |  |
| Q4 (least deprived) | 1 (ref) | ·· | 1 (ref) | ·· | 1 (ref) | ·· |
| Q3 | 0.71 (0.50–1.01) | 0.056 | 0.75 (0.51–1.12) | 0.161 | 0.86 (0.65–1.15) | 0.307 |
| Q2 | 1.00 (0.70–1.42) | 0.987 | 1.01 (0.67–1.51) | 0.972 | 0.98 (0.72–1.32) | 0.875 |
| Q1 (most deprived) | 0.98 (0.69–1.40) | 0.913 | 0.95 (0.63–1.42) | 0.796 | 1.08 (0.80–1.45) | 0.609 |
| BMI (kg/m²) | 1.02 (0.99–1.04) | 0.204 | 1.02 (1.00–1.05) | 0.098 | 1.02 (1.00–1.04) | 0.016 |
| General health |  |  |  |  |  |  |
| Excellent | 1 (ref) | ·· | 1 (ref) | ·· | 1 (ref) | ·· |
| Very good | 1.18 (0.81–1.71) | 0.398 | 1.56 (0.96–2.54) | 0.073 | 1.33 (0.97–1.83) | 0.081 |
| Good | 1.71 (1.13–2.58) | 0.011 | 2.56 (1.54–4.25) | <0.001 | 1.98 (1.40–2.80) | <0.001 |
| Fair | 2.86 (1.69–4.85) | <0.001 | 4.09 (2.21–7.57) | <0.001 | 3.07 (1.96–4.82) | <0.001 |
| Poor | 2.42 (0.98–6.02) | 0.056 | 2.50 (0.92–6.77) | 0.072 | 1.84 (0.81–4.16) | 0.144 |
| Number of comorbidities | 1.27 (1.08–1.48) | 0.003 | 1.28 (1.08–1.51) | 0.005 | 1.20 (1.05–1.36) | 0.007 |
| Any depression or anxiety (PHQ-4) | 1.01 (0.69–1.47) | 0.978 | 1.08 (0.72–1.63) | 0.705 | 1.15 (0.84–1.58) | 0.388 |
| Vigorous physical exercise, h per week |  |  |  |  |  |  |
| 0 | 1 (ref) | ·· | 1 (ref) | ·· | 1 (ref) | ·· |
| 1–3 | 0.80 (0.59–1.07) | 0.129 | 0.93 (0.67–1.31) | 0.689 | 0.97 (0.76–1.24) | 0.805 |
| ≥4 | 0.74 (0.52–1.07) | 0.106 | 0.97 (0.65–1.47) | 0.899 | 0.95 (0.71–1.29) | 0.763 |
| Self-reported infection severity |  |  |  |  |  |  |
| Asymptomatic | 1 (ref) | ·· | 1 (ref) | ·· | 1 (ref) | ·· |
| Mildly unwell | 1.41 (0.81–2.47) | 0.228 | 0.71 (0.37–1.40) | 0.328 | 1.25 (0.78–2.00) | 0.346 |
| Moderately unwell | 2.79 (1.62–4.80) | <0.001 | 2.33 (1.26–4.32) | 0.007 | 2.42 (1.53–3.82) | <0.001 |
| Very unwell | 5.33 (3.10–9.18) | <0.001 | 4.58 (2.49–8.43) | <0.001 | 4.35 (2.75–6.88) | <0.001 |
| Vaccinated at time of infection | 0.52 (0.35–0.78) | 0.002 | 0.53 (0.34–0.83) | 0.006 | 0.54 (0.39–0.76) | <0.001 |
| Variant |  |  |  |  |  |  |
| Pre-Omicron | 1 (ref) | ·· | 1 (ref) | ·· | 1 (ref) | ·· |
| Post-Omicron | 0.24 (0.17–0.35) | <0.001 | 0.30 (0.20–0.46) | <0.001 | 0.29 (0.21–0.39) | <0.001 |
| Baseline questionnaire | 1.03 (0.97–1.09) | 0.285 | 1.04 (0.97–1.11) | 0.284 | 1.04 (0.99–1.09) | 0.082 |
| Follow-up from SARS-CoV-2 infection, months | 1.04 (1.01–1.07) | 0.002 | 1.04 (1.01–1.07) | 0.007 | 1.04 (1.02–1.06) | <0.001 |

Sensitivity analysis 1 was restricted to participants with test-confirmed SARS-CoV-2 infection (3477 who did not report long COVID and 324 who did). Sensitivity analysis 2 restricted participants with long COVID to the 244 participants with a higher symptom burden (reporting at least six of the 16 symptoms investigated). The exploratory analysis extended the definition of long COVID to include the response “Don’t know / not sure” as a positive response, resulting in 483 participants reporting long COVID. BMI=body-mass index. IMD=Index of Multiple Deprivation. OR=odds ratio. PHQ-4=Patient Health Questionnaire-4.

### ***Table S4:* Regression estimates from logistic regression: sleep duration and sleep efficiency**

|  | **Model including sleep duration** | | **Model including sleep efficiency** | |
| --- | --- | --- | --- | --- |
|  | OR (95% CI) | p value | OR (95% CI) | p value |
| Sleep duration, hours |  |  |  |  |
| <5 | 1.75 (0.56–5.46) | 0.333 | ·· | ·· |
| 5–6 | 1.21 (0.90–1.61) | 0.210 | ·· | ·· |
| 7 | 1 (ref) | ·· | ·· | ·· |
| 8–9 | 0.77 (0.55–1.08) | 0.132 | ·· | ·· |
| >9 | 2.59 (0.62–10.85) | 0.194 | ·· | ·· |
| Sleep efficiency (per 10% increase) | ·· | ·· | 0.85 (0.76–0.95) | 0.004 |
| Sleep midpoint | 1.00 (0.90–1.12) | 0.865 | 0.99 (0.89–1.10) | 0.879 |
| SD of mid-sleep point (10 min) | 0.99 (0.96–1.02) | 0.431 | 0.99 (0.96–1.02) | 0.480 |
| SD of sleep efficiency (%) | 1.06 (1.01–1.11) | 0.012 | 1.07 (1.02–1.12) | 0.007 |
| SD of time asleep (10 min) | 1.00 (0.92–1.09) | 0.850 | 0.98 (0.90–1.07) | 0.636 |

ORs shown are from fully adjusted models. Although the point estimates suggest a quadratic trend in sleep duration, the trend test was not significant (p =0.095). OR=odds ratio.

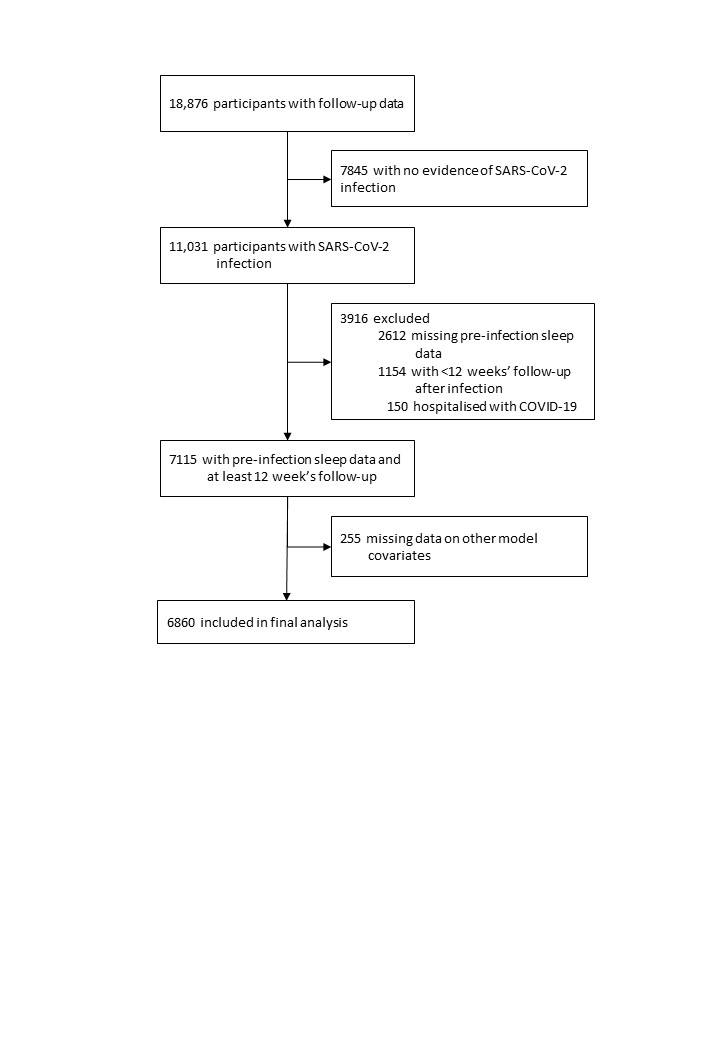

### ***Figure S5:* Participant flow diagram: analysis on post-infection sleep duration**

### ***Table S5:* Regression estimates from multilevel mixed model**

|  | **β (95% CI)** | **p value** |
| --- | --- | --- |
| COVID-19 status |  |  |
| No COVID | Reference | ·· |
| Infection ≤1 month prior | 0.028 (–0.007 to 0.064) | 0.113 |
| Infection >1 to 3 months prior | 0.005 (–0.017 to 0.027) | 0.679 |
| Infection >1 to 6 months prior | 0.005 (–0.017 to 0.028) | 0.637 |
| Infection >6 to 9 months prior | 0.004 (–0.022 to 0.030) | 0.760 |
| Infection >9 to 12 months prior | 0.031 (0.002 to 0.060) | 0.039 |
| Age, years | –0.005 (–0.007 to –0.003) | <0.001 |
| Sex |  |  |
| Female | 0.029 (–0.010 to 0.067) | 0.143 |
| Male | Reference | ·· |
| Highest educational level attained |  |  |
| Primary or secondary | –0.059 (–0.121 to 0.003) | 0.063 |
| Higher or further (A levels) | –0.030 (–0.084 to 0.024) | 0.278 |
| College or university | –0.011 (–0.050 to 0.028) | 0.580 |
| Post-graduate | Reference | ·· |
| Quartiles of IMD decile |  |  |
| Q4 (least deprived) | Reference | ·· |
| Q3 | 0.028 (–0.015 to 0.071) | 0.207 |
| Q2 | 0.029 (–0.018 to 0.076) | 0.229 |
| Q1 (most deprived) | –0.060 (–0.109 to –0.011) | 0.016 |
| BMI (kg/m²) | –0.005 (–0.008 to –0.001) | 0.009 |
| General health |  |  |
| Excellent | Reference | ·· |
| Very good | –0.019 (–0.065 to 0.026) | 0.403 |
| Good | –0.039 (–0.091 to 0.013) | 0.145 |
| Fair | 0.030 (–0.042 to 0.103) | 0.411 |
| Poor | 0.044 (–0.087 to 0.175) | 0.512 |
| Number of comorbidities | –0.005 (–0.025 to 0.015) | 0.623 |
| Any SARS-CoV-2 vaccinations | –0.009 (–0.028 to 0.009) | 0.326 |
| Baseline sleep problems | –0.279 (–0.319 to –0.240) | <0.001 |
| Baseline sleep quality |  |  |
| Good | Reference | ·· |
| Medium | –0.385 (–0.426 to –0.345) | <0.001 |
| Medium–low | –0.800 (–0.850 to –0.751) | <0.001 |
| Low | –1.595 (–1.681 to –1.509) | <0.001 |
| Baseline questionnaire |  |  |
| Oct 2020 | Reference | ·· |
| Nov 2020 | –0.041 (–0.079 to –0.004) | 0.028 |
| Dec 2020 | 0.046 (–0.049 to 0.141) | 0.339 |
| Jan 2021 | 0.102 (–0.040 to 0.243) | 0.159 |
| Feb 2021 | –0.067 (–0.240 to 0.107) | 0.451 |
| March 2021 | –0.358 (–0.606 to –0.109) | 0.005 |
| April 2021 | –0.010 (–0.145 to 0.125) | 0.886 |
| May 2021 | –0.048 (–0.272 to 0.176) | 0.674 |
| Occupational status |  |  |
| Employed/working | Reference | ·· |
| Not employed/not working | 0.116 (0.096 to 0.136) | <0.001 |
| Other | 0.088 (0.041 to 0.134) | <0.001 |
| Vigorous physical exercise, h per week |  |  |
| 0 | Reference | ·· |
| 1–3 | 0.022 (0.014 to 0.029) | <0.001 |
| ≥4 | 0.051 (0.041 to 0.061) | <0.001 |
| PHQ-4 grade |  |  |
| Normal | Reference | ·· |
| Mild | –0.113 (–0.124 to –0.102) | <0.001 |
| Moderate | –0.263 (–0.285 to –0.241) | <0.001 |
| Severe | –0.413 (–0.447 to –0.379) | <0.001 |
| Month/year of survey | –0.005 (–0.007 to –0.004) | <0.001 |
| Infection severity |  |  |
| Asymptomatic or mild | Reference | ·· |
| Moderately unwell | 0.033 (–0.008 to 0.074) | 0.114 |
| Very unwell | 0.113 (0.070 to 0.156) | <0.001 |
| Long COVID status (sleep problems) |  |  |
| Does not report long COVID | Reference | ·· |
| Long COVID: no reported sleep problems | –0.006 (–0.074 to 0.061) | 0.852 |
| Long COVID: reported sleep problems | –0.030 (–0.092 to 0.031) | 0.334 |
| Interaction between COVID-19 status and infection severity |  |  |
| Infection ≤1 month prior |  |  |
| Asymptomatic or mild | Reference | ·· |
| Moderately unwell | 0.072 (0.021 to 0.123) | 0.005 |
| Very unwell | 0.204 (0.151 to 0.257) | <0.001 |
| Infection >1 to 3 months prior |  |  |
| Asymptomatic or mild | Reference | ·· |
| Moderately unwell | –0.008 (–0.037 to 0.022) | 0.612 |
| Very unwell | 0.016 (–0.015 to 0.047) | 0.300 |
| Infection >3 to 6 months prior |  |  |
| Asymptomatic or mild | Reference | ·· |
| Moderately unwell | –0.023 (–0.051 to 0.005) | 0.111 |
| Very unwell | –0.013 (–0.042 to 0.016) | 0.371 |
| Infection >6 to 9 months prior |  |  |
| Asymptomatic or mild | Reference | ·· |
| Moderately unwell | –0.026 (–0.057 to 0.004) | 0.093 |
| Very unwell | –0.009 (–0.041 to 0.023) | 0.570 |
| Infection >9 to 12 months prior |  |  |
| Asymptomatic or mild | Reference | ·· |
| Moderately unwell | –0.045 (–0.079 to –0.010) | 0.012 |
| Very unwell | –0.029 (–0.066 to 0.008) | 0.122 |
| Interaction between COVID-19 status and long COVID status |  |  |
| Infection ≤1 month prior |  |  |
| Does not report long COVID | Reference | ·· |
| Long COVID: no reported sleep problems | 0.242 (0.063 to 0.420) | 0.008 |
| Long COVID: reported sleep problems | –0.183 (–0.468 to 0.102) | 0.209 |
| Infection >1 to 3 months prior |  |  |
| Does not report long COVID | Reference | ·· |
| Long COVID: no reported sleep problems | 0.099 (0.007 to 0.192) | 0.035 |
| Long COVID: reported sleep problems | –0.172 (–0.269 to –0.074) | <0.001 |
| Infection >3 to 6 months prior |  |  |
| Does not report long COVID | Reference | ·· |
| Long COVID: no reported sleep problems | 0.088 (0.003 to 0.173) | 0.041 |
| Long COVID: reported sleep problems | –0.097 (–0.192 to –0.002) | 0.046 |
| Infection >6 to 9 months prior |  |  |
| Does not report long COVID | Reference | ·· |
| Long COVID: no reported sleep problems | 0.051 (–0.040 to 0.143) | 0.273 |
| Long COVID: reported sleep problems | 0.019 (–0.107 to 0.145) | 0.769 |
| Infection >9 to 12 months prior |  |  |
| Does not report long COVID | Reference | ·· |
| Long COVID: no reported sleep problems | 0.105 (0.004 to 0.206) | 0.042 |
| Long COVID: reported sleep problems | –0.130 (–0.295 to 0.036) | 0.125 |

BMI=body-mass index.

### ***Table S6:* Contrasts of predictive margins**

|  | **Main analysis** | **Sensitivity 1** | **Sensitivity 2** |
| --- | --- | --- | --- |
| **Overall** |  |  |  |
| ≤1 month | 0.11 (0.09 to 0.14) | 0.12 (0.09 to 0.14) | 0.12 (0.09 to 0.15) |
| >1 to 3 months | 0.01 (–0.01 to 0.02) | 0.01 (–0.01 to 0.02) | 0.01 (–0.01 to 0.03) |
| >3 to 6 months | –0.01 (–0.02 to 0.01) | 0.00 (–0.02 to 0.02) | 0.00 (–0.02 to 0.02) |
| >6 to 9 months | –0.01 (–0.03 to 0.01) | 0.00 (–0.02 to 0.02) | 0.00 (–0.03 to 0.02) |
| >9 to 12 months | 0.01 (–0.02 to 0.03) | 0.01 (–0.01 to 0.03) | 0.01 (–0.02 to 0.04) |
| **By severity** |  |  |  |
| Asymptomatic or mild |  |  |  |
| ≤1 month | 0.03 (–0.01 to 0.07) | 0.03 (0.00 to 0.07) | 0.04 (0.00 to 0.09) |
| >1 to 3 months | 0.00 (–0.02 to 0.03) | 0.01 (–0.01 to 0.03) | 0.00 (–0.03 to 0.03) |
| >3 to 6 months | 0.01 (–0.02 to 0.03) | 0.01 (–0.01 to 0.04) | 0.01 (–0.02 to 0.04) |
| >6 to 9 months | 0.00 (–0.02 to 0.03) | 0.01 (–0.02 to 0.04) | 0.01 (–0.03 to 0.04) |
| >9 to 12 months | 0.03 (0.00 to 0.06) | 0.04 (0.01 to 0.07) | 0.03 (0.00 to 0.07) |
| Moderately unwell |  |  |  |
| ≤1 month | 0.10 (0.06 to 0.14) | 0.10 (0.06 to 0.14) | 0.10 (0.05 to 0.15) |
| >1 to 3 months | 0.00 (–0.03 to 0.02) | 0.00 (–0.03 to 0.02) | 0.00 (–0.03 to 0.03) |
| >3 to 6 months | –0.02 (–0.04 to 0.01) | –0.01 (–0.04 to 0.01) | –0.02 (–0.05 to 0.01) |
| >6 to 9 months | –0.02 (–0.05 to 0.01) | –0.02 (–0.04 to 0.01) | –0.02 (–0.05 to 0.02) |
| >9 to 12 months | –0.01 (–0.05 to 0.02) | –0.01 (–0.05 to 0.02) | –0.02 (–0.06 to 0.03) |
| Very unwell |  |  |  |
| ≤1 month | 0.23 (0.19 to 0.27) | 0.24 (0.20 to 0.28) | 0.23 (0.18 to 0.29) |
| >1 to 3 months | 0.02 (–0.01 to 0.04) | 0.02 (–0.01 to 0.04) | 0.02 (–0.01 to 0.05) |
| >3 to 6 months | –0.01 (–0.03 to 0.02) | –0.01 (–0.03 to 0.02) | 0.00 (–0.03 to 0.03) |
| >6 to 9 months | 0.00 (–0.03 to 0.03) | 0.00 (–0.03 to 0.03) | 0.01 (–0.03 to 0.04) |
| >9 to 12 months | 0.00 (–0.03 to 0.04) | 0.00 (–0.03 to 0.04) | 0.01 (–0.04 to 0.05) |
| **By long COVID status** |  |  |  |
| Does not report long COVID |  |  |  |
| ≤1 month | 0.11 (0.09 to 0.13) | 0.12 (0.09 to 0.14) | 0.12 (0.09 to 0.15) |
| >1 to 3 months | 0.01 (–0.01 to 0.02) | 0.01 (–0.01 to 0.02) | 0.01 (–0.01 to 0.03) |
| >3 to 6 months | –0.01 (–0.02 to 0.01) | 0.00 (–0.02 to 0.02) | 0.00 (–0.02 to 0.02) |
| >6 to 9 months | –0.01 (–0.03 to 0.01) | 0.00 (–0.02 to 0.02) | 0.00 (–0.03 to 0.02) |
| >9 to 12 months | 0.01 (–0.02 to 0.03) | 0.01 (–0.02 to 0.03) | 0.01 (–0.02 to 0.04) |
| Reports long COVID without sleep problems |  |  |  |
| ≤1 month | 0.38 (0.20 to 0.56) | 0.38 (0.20 to 0.56) | 0.13 (–0.10 to 0.37) |
| >1 to 3 months | 0.11 (0.02 to 0.20) | 0.11 (0.02 to 0.20) | 0.07 (–0.04 to 0.18) |
| >3 to 6 months | 0.08 (0.00 to 0.17) | 0.08 (0.00 to 0.17) | 0.07 (–0.04 to 0.18) |
| >6 to 9 months | 0.04 (–0.05 to 0.13) | 0.05 (–0.04 to 0.14) | 0.00 (–0.11 to 0.12) |
| >9 to 12 months | 0.11 (0.01 to 0.21) | 0.11 (0.01 to 0.21) | 0.10 (–0.02 to 0.23) |
| Reports long COVID with sleep problems |  |  |  |
| ≤1 month | –0.03 (–0.31 to 0.26) | –0.03 (–0.31 to 0.26) | 0.08 (–0.26 to 0.42) |
| >1 to 3 months | –0.16 (–0.26 to –0.06) | –0.16 (–0.26 to –0.06) | –0.36 (–0.49 to –0.23) |
| >3 to 6 months | –0.10 (–0.20 to –0.01) | –0.10 (–0.19 to 0.00) | –0.16 (–0.28 to –0.04) |
| >6 to 9 months | 0.01 (–0.11 to 0.14) | 0.01 (–0.11 to 0.14) | –0.01 (–0.17 to 0.16) |
| >9 to 12 months | –0.12 (–0.29 to 0.04) | –0.16 (–0.32 to 0.01) | –0.33 (–0.55 to –0.11) |

Sensitivity analysis 1 was restricted to participants with test-confirmed SARS-CoV-2 infection (n=6476). Sensitivity analysis 2 was restricted to participants with pre-infection sleep data from October, 2020 (n=4216).

### **Selection bias and missing data**

#### *Long COVID risk analysis*

Among all participants reporting SARS-CoV-2 infection (n=11,031), key characteristics were compared between those included in the analysis and those excluded. Excluded participants were younger, had lower educational attainment, did less physical exercise, had higher BMI, poorer general health, and a greater prevalence of comorbidities. Those with pre-infection sleep data were more likely to have poorer quality sleep and lower sleep efficiency. Excluded participants were also more likely to be infected before dominance of the Omicron variant, less likely to have been vaccinated when infected, and experienced more severe disease.

#### ***Table S7:* Comparison of key characteristics between participants included and excluded from the long COVID risk analysis**

|  | **Excluded (N=7037)** | **Included (N=3994)** |
| --- | --- | --- |
| **Sociodemographic and behavioural** |  |  |
| Age, years | 59.6 (49.8–67.4) | 63.1 (54.8–69.1) |
| Sex |  |  |
| Female | 5150 (73.2%) | 2844 (71.2%) |
| Male | 1887 (26.8%) | 1150 (28.8%) |
| Ethnicity |  |  |
| White | 6636 (94.3%) | 3853 (96.5%) |
| Black, African, Caribbean, or Black British | 44 (0.6%) | 20 (0.5%) |
| South Asian | 135 (1.9%) | 51 (1.3%) |
| Mixed, multiple, or other | 221 (3.1%) | 70 (1.8%) |
| IMD decile | 7 (5–9) | 7 (5–9) |
| Highest educational level attained |  |  |
| Primary or secondary | 712/7026 (10.1%) | 379 (9.5%) |
| Higher or further (A levels) | 999/7026 (14.2%) | 537 (13.4%) |
| College or university | 3165/7026 (45.0%) | 1789 (44.8%) |
| Post-graduate | 2150/7026 (30.6%) | 1289 (32.3%) |
| Weekly vigorous physical exercise |  |  |
| 0 h | 1670/4560 (36.6%) | 1255 (31.4%) |
| 1–3 h | 1959/4560 (43.0%) | 1810 (45.3%) |
| ≥4 h | 931/4560 (20.4%) | 929 (23.3%) |
| **Baseline sleep characteristics** |  |  |
| Sleep duration, h | 6.9 (1.1) | 6.9 (1.0) |
| Sleep efficiency | 82.0 (74.0–90.0) | 84.0 (75.0–90.0) |
| Sleep quality |  |  |
| Good | 2,204/6538 (33.7%) | 1549 (38.8%) |
| Medium | 2,505/6538 (38.3%) | 1592 (39.9%) |
| Medium–low | 1,429/6538 (21.9%) | 747 (18.7%) |
| Low | 400/6538 (6.1%) | 106 (2.7%) |
| **Clinical characteristics** |  |  |
| BMI, kg/m² |  |  |
| <25 | 3181/7013 (45.4%) | 2127 (53.3%) |
| 25 to <30 | 2289/7013 (32.6%) | 1245 (31.2%) |
| ≥30 | 1543/7013 (22.0%) | 622 (15.6%) |
| General health |  |  |
| Excellent | 1040/7034 (14.8%) | 1049 (26.3%) |
| Very good | 2460/7034 (35.0%) | 1777 (44.5%) |
| Good | 2062/7034 (29.3%) | 887 (22.2%) |
| Fair | 1036/7034 (14.7%) | 231 (5.8%) |
| Poor | 436/7034 (6.2%) | 50 (1.3%) |
| Asthma or COPD | 1488 (21.1%) | 585 (14.6%) |
| Autoimmune disease | 696 (9.9%) | 323 (8.1%) |
| Diabetes |  |  |
| Pre-diabetes | 223 (3.2%) | 114 (2.9%) |
| Diabetes | 325 (4.6%) | 164 (4.1%) |
| Heart disease | 271 (3.9%) | 116 (2.9%) |
| Hypertension | 1476 (21.0%) | 827 (20.7%) |
| Immunosuppression or organ transplant | 395 (5.6%) | 152 (3.8%) |
| PHQ-4 grade |  |  |
| Normal | 5286/7011 (75.4%) | 3522 (88.3%) |
| Mild | 1200/7011 (17.1%) | 372 (9.3%) |
| Moderate | 350/7011 (5.0%) | 67 (1.7%) |
| Severe | 175/7011 (2.5%) | 29 (0.7%) |
| **Acute infection and vaccination status** |  |  |
| Likely COVID variant |  |  |
| Pre-Omicron | 3709 (52.7%) | 747 (18.7%) |
| Omicron | 3328 (47.3%) | 3247 (81.3%) |
| Self-reported infection severity |  |  |
| Asymptomatic | 271/5534 (4.9%) | 280 (7.0%) |
| Mildly unwell | 1331/5534 (24.1%) | 1465 (36.7%) |
| Moderately unwell | 1631/5534 (29.5%) | 1313 (32.9%) |
| Very unwell | 2301/5534 (41.6%) | 935 (23.4%) |
| Vaccinated at time of infection | 3897 (55.4%) | 3672 (91.9%) |

The table below shows the pattern of missing covariate data, among the 106 participants missing data on either educational attainment (edu), Index of Multiple Deprivation (IMD), body-mass index (BMI), or infection severity (sev). The vast majority (88%) were missing data on infection severity alone.

|  | **edu** | **IMD** | **BMI** | **sev** |
| --- | --- | --- | --- | --- |
| 88% |  |  |  |  |
| 6% |  |  |  |  |
| 5% |  |  |  |  |
| 2% |  |  |  |  |

#### *Post-infection sleep analysis*

Among all participants reporting SARS-CoV-2 infection (n=11,031), key characteristics were compared between those included in the analysis and those excluded. Excluded participants were younger, less likely to be White, had higher BMI, poorer general health, and a greater prevalence of asthma or COPD. Those with pre-infection sleep data were more likely to have poorer quality sleep. Excluded participants were also less likely to have been vaccinated when infected, and experienced more severe disease.

#### ***Table S8:* Comparison of key characteristics between participants included and excluded from the post-infection sleep analysis**

|  | **Excluded (N=4171)** | **Included (N=6860)** |
| --- | --- | --- |
| **Sociodemographic and behavioural** |  |  |
| Age, years | 57.2 (47.5–65.6) | 63.0 (54.2–69.1) |
| Sex |  |  |
| Female | 3096 (74.2%) | 4898 (71.4%) |
| Male | 1075 (25.8%) | 1962 (28.6%) |
| Ethnicity |  |  |
| White | 3894 (93.4%) | 6595 (96.1%) |
| Black, African, Caribbean, or Black British | 36 (0.9%) | 28 (0.4%) |
| South Asian | 86 (2.1%) | 100 (1.5%) |
| Mixed, multiple, or other | 154 (3.7%) | 137 (2.0%) |
| IMD decile | 7 (4–9) | 7 (5–9) |
| Highest educational level attained |  |  |
| Primary or secondary | 420/4160 (10.1%) | 671 (9.8%) |
| Higher or further (A levels) | 580/4160 (13.9%) | 956 (13.9%) |
| College or university | 1861/4160 (44.7%) | 3093 (45.1%) |
| Post-graduate | 1299/4160 (31.2%) | 2140 (31.2%) |
| **Baseline sleep characteristics*** |  |  |
| Sleep duration, h | 6.9 (1.2) | 6.9 (1.0) |
| Sleep quality |  |  |
| Good | 426/1330 (32.0%) | 2290 (33.4%) |
| Medium | 520/1330 (39.1%) | 2731 (39.8%) |
| Medium–low | 275/1330 (20.7%) | 1508 (22.0%) |
| Low | 109/1330 (8.2%) | 331 (4.8%) |
| Sleep problems | 1079/3025 (35.7%) | 2247 (32.8%) |
| **Clinical characteristics** |  |  |
| BMI, kg/m² |  |  |
| <25 | 1892/4147 (45.6%) | 3416 (49.8%) |
| 25 to <30 | 1326/4147 (32.0%) | 2208 (32.2%) |
| ≥30 | 929/4147 (22.4%) | 1236 (18.0%) |
| General health |  |  |
| Excellent | 643/4168 (15.4%) | 1446 (21.1%) |
| Very good | 1382/4168 (33.2%) | 2855 (41.6%) |
| Good | 1155/4168 (27.7%) | 1794 (26.2%) |
| Fair | 636/4168 (15.3%) | 631 (9.2%) |
| Poor | 352/4168 (8.4%) | 134 (2.0%) |
| Asthma or COPD | 917 (22.0%) | 1156 (16.9%) |
| Autoimmune disease | 408 (9.8%) | 611 (8.9%) |
| Diabetes |  |  |
| Pre-diabetes | 125 (3.0%) | 212 (3.1%) |
| Diabetes | 189 (4.5%) | 300 (4.4%) |
| Heart disease | 128 (3.1%) | 259 (3.8%) |
| Hypertension | 801 (19.2%) | 1502 (21.9%) |
| Immunosuppression or organ transplant | 225 (5.4%) | 322 (4.7%) |
| PHQ-4 grade |  |  |
| Normal | 3005/4155 (72.3%) | 5803 (84.8%) |
| Mild | 781/4155 (18.8%) | 791 (11.6%) |
| Moderate | 247/4155 (5.9%) | 170 (2.5%) |
| Severe | 122/4155 (2.9%) | 82 (1.2%) |
| **Acute infection and vaccination status** |  |  |
| Self-reported infection severity |  |  |
| Asymptomatic | 106/2704 (3.9%) | 445 (6.5%) |
| Mildly unwell | 638/2704 (23.6%) | 2171 (31.6%) |
| Moderately unwell | 714/2704 (26.4%) | 2244 (32.7%) |
| Very unwell | 1246/2704 (46.1%) | 2000 (29.2%) |
| Vaccinated at time of infection | 2838 (68.0%) | 5456 (79.5%) |
